# GLOW UP Study: Protocol for an Observational Digital Biomarker Study for Prediabetes Screening and Digital Phenotyping

**DOI:** 10.64898/2026.03.11.26348172

**Authors:** Victoria Brügger, Magdalena Fuchs, Qiuhan Jin, Benjamin Wirth, Stefan Bilz, Michael Brändle, Tobias Kowatsch, Mia Jovanova

**Affiliations:** Digital Health Interventions, School of Medicine, University of St. Gallen, St. Gallen, Switzerland; Centre for Digital Health Interventions, Department of Management, Technology and Economics, ETH Zürich, Zurich, Switzerland; Department of Endocrinology, HOCH Health Ostschweiz, St. Gallen, Switzerland; Department of Internal Medicine, HOCH Health Ostschweiz, St. Gallen, Switzerland; Digital Health Interventions, Institute for Implementation Science in Health Care, University of Zurich, Zurich, Switzerland

**Keywords:** digital phenotyping, digital biomarkers, diabetes, prevention, lifestyle

## Abstract

**Introduction:** Prediabetes, a key precursor to type 2 diabetes, is highly prevalent and underdiagnosed, particularly among adults aged ≥45 years with elevated body mass index (BMI). Early detection is critical because lifestyle interventions can delay or prevent progression to type 2 diabetes. The Glow Up (GLucose Observation and Wearable Use for Prevention) study aims to (1) test the feasibility of a digital biomarker for prediabetes screening using wearable– and smartphone-derived lifestyle factors (e.g., sleep, physical activity, and nutrition patterns), in daily life and to (2) characterize individual– and metabolic subgroup-level variability in lifestyle factors and glycemic control. Specifically, we aim to examine how lifestyle factors relate to diabetes risk and identify personalized predictors of early metabolic dysregulation.

**Methods and analysis:** Glow Up is a prospective, single-center, observational case-control study conducted in Switzerland. Adults (N=200) aged ≥45 years with BMI ≥25 kg/m² will be recruited, including n=100 individuals with prediabetes and n=100 age– and sex-matched case-control normoglycemic controls. Participants will undergo four weeks of continuous monitoring using a blinded continuous glucose monitor (CGM), and commercial– and medical-grade wearables; e.g. capturing physical activity, sleep, and physiological markers (heart rate variability, heart rate and skin temperature); in addition to completing daily image-based meal logs, using smartphones. Glycated haemoglobin (HbA1c), fasting plasma glucose (FPG) and body anthropometrics will be collected at baseline and follow-up, four weeks apart. Primary outcomes include HbA1c and FPG, measured at approximately four week follow-up. Secondary outcomes include CGM metrics, lifestyle profiling (sleep, physical activity, stress, and nutrition), and adherence to image-based meal logging.

**Ethics and dissemination:** The study has received ethics approval from the Ethics Committee of Eastern Switzerland (BASEC ID.: 2025-00972). Results will be published in international peer-reviewed journals and at national and international conferences as posters, presentations, and articles. Summaries will be provided to the funders and personalized reports to participants.

**Trial registration number:** NCT07373418

**Article Summary:** *Strengths and limitations of this study:*

- Glow Up presents the first study in Switzerland to investigate glycaemic and lifestyle profiling among individuals with normoglycaemia and prediabetes in an observational, free-living setting.
- To our knowledge, Glow Up is the first dataset to combine continuous glucose monitoring, commercial– and medical-grade wearable sensors, smartphone-based questionnaires, image-based dietary logging under free-living conditions, and clinical biomarkers, HbA1c and FPG; allowing for rich digital phenotyping.
- Inclusion of baseline and follow-up clinical biomarkers (HbA1c and FPG) provides clinically validated ground truth measures, strengthening outcome validity and anchoring digital and wearable-derived markers to established clinical, diagnostic standards.
- The absence of direct measures of insulin resistance, lipid metabolism, and gut microbiome compositions may limit more mechanistic investigation of glucose dysregulation patterns and metabolic sub phenotyping.
- The four-week follow-up period may limit inference on longer-term glycaemic trajectories and progression to T2D.
- Findings may not be generalisable beyond at-risk adults living in Switzerland who meet the study eligibility criteria.

## INTRODUCTION

Diabetes presents a major public health and economic challenge, with significant global and national implications. In Switzerland, around half a million people live with diabetes, 90% of which is attributed to Type 2 Diabetes (T2D), incurring direct healthcare costs of 1.7 billion CHF annually (1). Globally, the prevalence of T2D is expected to double to 1.2 billion by 2050 (2). Prediabetes, a condition characterized by elevated blood glucose levels, is a key risk factor for the development of T2D, as well as for cardiovascular disease and stroke (3–5). Each year, approximately 5-10 % of individuals with prediabetes progress to T2D (6). Early detection of prediabetes is thus critical to enable timely lifestyle interventions that can prevent or revert T2D, as well as its complications, and reduce associated direct costs (7,8). However, undetected prediabetes prevalence remains high (9). In Switzerland, a study of adults aged 60 and older (N=1’362) found that 8.4% (n=114) had undiagnosed T2D, while 64.5% (n=878) had undiagnosed prediabetes (3); highlighting the high levels of prediabetes in the key at-risk population. Globally, an estimated 240 million people are living with undiagnosed diabetes (10), highlighting the need for innovative screening methods to identify at-risk individuals earlier; understand metabolic dysregulation patterns, and deliver more targeted interventions (11).

Diabetes and prediabetes are clinically diagnosed through blood tests, such as glycated haemoglobin (HbA1c) and fasting plasma glucose (FPG). HbA1c reflects average blood sugar levels over the past three months (12), while FPG measures glucose at a single point in time; typically after an 8–12 hour overnight fast and primarily captures short-term glucose regulation driven by hepatic glucose production. Following standard clinical thresholds, individuals with HbA1c <5.7% (39 mmol/mol) and FPG <5.6 mmol/L are classified as normoglycemic (HbA1c-/FPG-); HbA1c 5.7-6.4% (39–46 mmol/mol) or FPG 5.6-6.9 mmol/L as prediabetic; HbA1c ≥6.5% (48 mmol/mol) or FPG ≥7.0 mmol/L, as T2D (13). In Switzerland, FPG and HbA1c testing is generally recommended every three to five years for at-risk individuals, particularly those over 45, with BMI ≥ 25 kg/m², a family history of T2D, and a sedentary lifestyle (14). However, standard blood tests may miss preclinical metabolic changes that precede HbA1c and FPG diagnostic thresholds, particularly over 3-5 year long screening intervals (7). Furthermore, many individuals at-risk of T2D remain untested, possibly in part due to limited awareness of the importance of diagnostic blood tests, potential barriers to healthcare access, and lack of early symptoms (15). Together, these factors likely contribute to the underdiagnosis of prediabetes and diabetes, delaying intervention, and increasing the likelihood of T2D related health complications (16).

In parallel, emerging research suggests that physiological signals captured by medical-grade wearable sensors; particularly electrocardiogram (ECG)-derived metrics such as heart rate variability (HRV); may provide complementary markers of early metabolic dysregulation. Consumer-grade wearables equipped with photoplethysmography (PPG) sensors are increasingly used to measure cardiovascular parameters, and many studies have evaluated their validity and accuracy compared with clinical reference methods (17–19). Dysglycemia and insulin resistance are associated with autonomic nervous system dysfunction and alterations in cardiac electrophysiology, which can be reflected in ECG patterns (20,21). Differences in ECG patterns between individuals with and without diabetes have enabled successful classification of disease status, with classification accuracies of 86.9% (22) and 90% (23). Similarly, PPG-derived metrics, which capture cardiovascular dynamics, have shown promise for diabetes detection. PPG measures can distinguish diabetic from normoglycemic individuals with area-under-the-curve (AUC) values ranging from 69.4% (24) to 83.0% (25), and can further differentiate normoglycemia, prediabetes, and T2D with an accuracy of 84% relative to ground truth blood test measures (26). While these findings highlight the potential of ECG and prototype PPG sensors as indirect indicators of metabolic dysregulation, most evidence to date has been generated in controlled or semi-controlled settings, using specialized devices. As a result, their translation into scalable, real-world screening in daily life remains limited.

In contrast to ECG devices, smartphones are widely ubiquitous, with wearable devices like smartwatches and smart rings becoming increasingly popular. The growing adoption of smartphones, smartwatches and smart rings, provide new opportunities to continuously track lifestyle behaviors, including activity, sleep, and heart rate, that are associated with metabolic function (27). Importantly, clinical studies have shown that adverse lifestyle factors are associated with an increased risk of T2D: for example, lower level of physical activity (28,29), insufficient sleep duration (30), and high intake of refined carbohydrates (31,32) have been linked to a higher risk of developing T2D and are more prevalent among individuals with diabetes compared to individuals with prediabetes or normoglycemia (33). These findings raise the question of whether continuous monitoring of lifestyle behaviors using commercially available digital devices could serve as an early indicator of metabolic dysregulation, potentially enabling detection of prediabetes before traditional blood tests. Specifically, smartphone usage among middle-aged and older adults in Switzerland has increased significantly in recent years. A 2020 study found that 74% of individuals aged 65 and above use web apps, with mobile web usage (via smartphones or tablets) in this age group surging from 31% in 2014 to 68% in 2020 (34). Furthermore, 50% of adults in Switzerland use ‘lifelogging’ applications to track daily activities on their smartphones (35). In parallel, wearable devices, such as Garmin smartwatches, are gaining popularity among middle-aged and older adults. In 2020, only 4.4% to 6.6% owned a smartwatch (36), but by 2023, usage had nearly doubled to 12%, with most users wearing them daily (37). In the same year approximately, more than one third of the population used personal connected devices such as smartwatches, smart rings, fitness bands, glasses, GPS trackers, or similar devices (37). The widespread use of smartphones and the growing adoption of fitness trackers, such as smartwatches and smart rings, suggest that these devices could enable early T2D risk screening and provide insights into individual lifestyle and glycaemic patterns (38). However, to our knowledge, no study in Switzerland has yet evaluated whether lifestyle data from commercially available devices can reliably distinguish prediabetes from normoglycemia in daily life.

### Objectives of the Study

To address this gap, the Glow Up study aims to evaluate the feasibility of a new digital biomarker for prediabetes screening in daily life. The primary objective is to examine whether, and to what extent, wearable and smartphone collected lifestyle data, e.g., a combination of sleep, physical activity, nutrition, heart rate (HR) and HRV over approximately four weeks, can distinguish the lifestyle patterns of individuals with prediabetes from normoglycemic controls, using ground truth HbA1c and FPG as primary outcomes. Secondary objectives include characterizing digital phenotypes at both the individual and metabolic subgroup levels. Specifically, we aim to assess individual-level variability by examining how lifestyle profiles contribute to time-varying continuous glucose monitoring (CGM)-derived metrics (i.e. postprandial glucose), and to evaluate how digital phenotypes, including lifestyle behaviors and glycemic patterns, differ across metabolic subgroups (e.g., defined by fasting plasma glucose, HbA1c, BMI, and visceral fat). We will also test image-based nutrition composition on glycemic control and evaluate adherence to a ∼4-week image-based meal logging protocol, including the impact of smartphone-based reminder messages on meal-logging adherence (39). See Table A1 for overview of primary and secondary outcomes of the study.

## METHODS

This protocol follows the Standardized Protocol Items Recommendations for Observational Studies (SPIROS) (see Appendix Table A3).

### Study design and setting

The Glow Up study is a prospective, single-center, observational case–control study conducted in Switzerland, enrolling 200 adults aged ≥45 years with BMI ≥25 kg/m², including 100 individuals with prediabetes and 100 age– and sex-matched normoglycemic controls. The study combines on-site visits with approximately up to ∼four weeks of remote lifestyle monitoring. Participants complete four visits: enrolment and eligibility (T0), baseline wearable onboarding (T1), intermediate device exchange (T2), and follow-up with wearable offboarding and blood tests (T3). At the baseline visit (T1), participants are provided with a smartwatch, smart ring, a blinded continuous glucose monitor, and a smartphone application for image-based meal logging. Baseline FPG and HbA1c are measured at baseline visit (T1) and repeated at the follow-up visit (T3), approximately four weeks later. Refer to Figure 1 for the study timeline.

**Figure 1.**
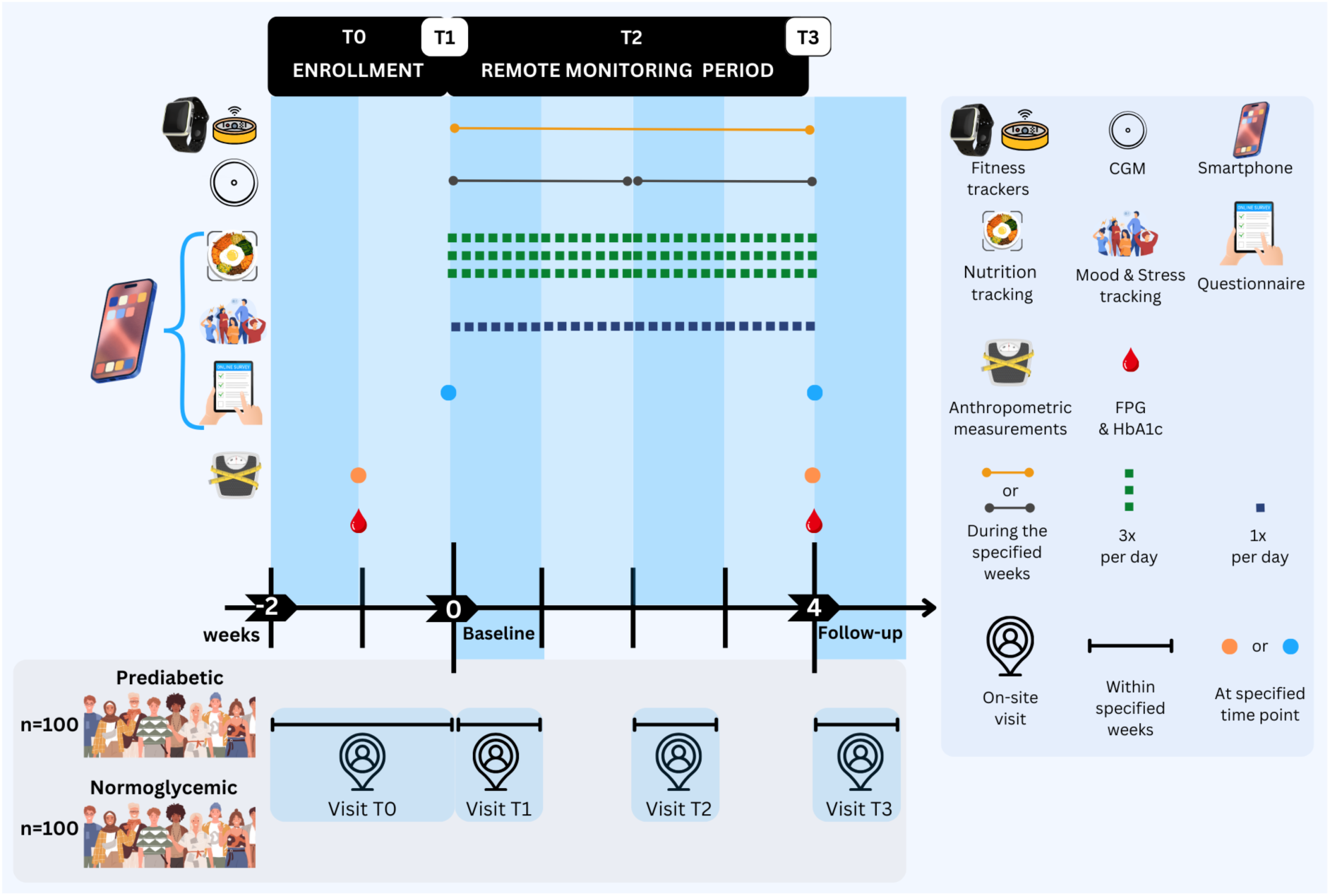
Study Timeline. The study consists of four stages: recruitment and enrolment (T0), during which written consent will be obtained, a baseline visit (T1), a four-week remote monitoring period with an intermediate visit for device exchange (T2), and a follow-up on-site visit (T3). Note: For illustrative purposes, continuous glucose monitoring (CGM), fasting plasma glucose (FPG), and haemoglobin A1c (HbA1c) are abbreviated.

### Study procedure

#### Participants, recruitment and screening

The study recruits two cohorts: adults at-risk of T2D (40) based on BMI and age (BMI ≥25 kg/m² and age 45), with prediabetes cases more likely to progress to T2D (41), and individuals who meet FPG and/or HbA1c diagnostic criteria for prediabetes. The eligibility criteria are summarized in Table 1.

**Table 1.**
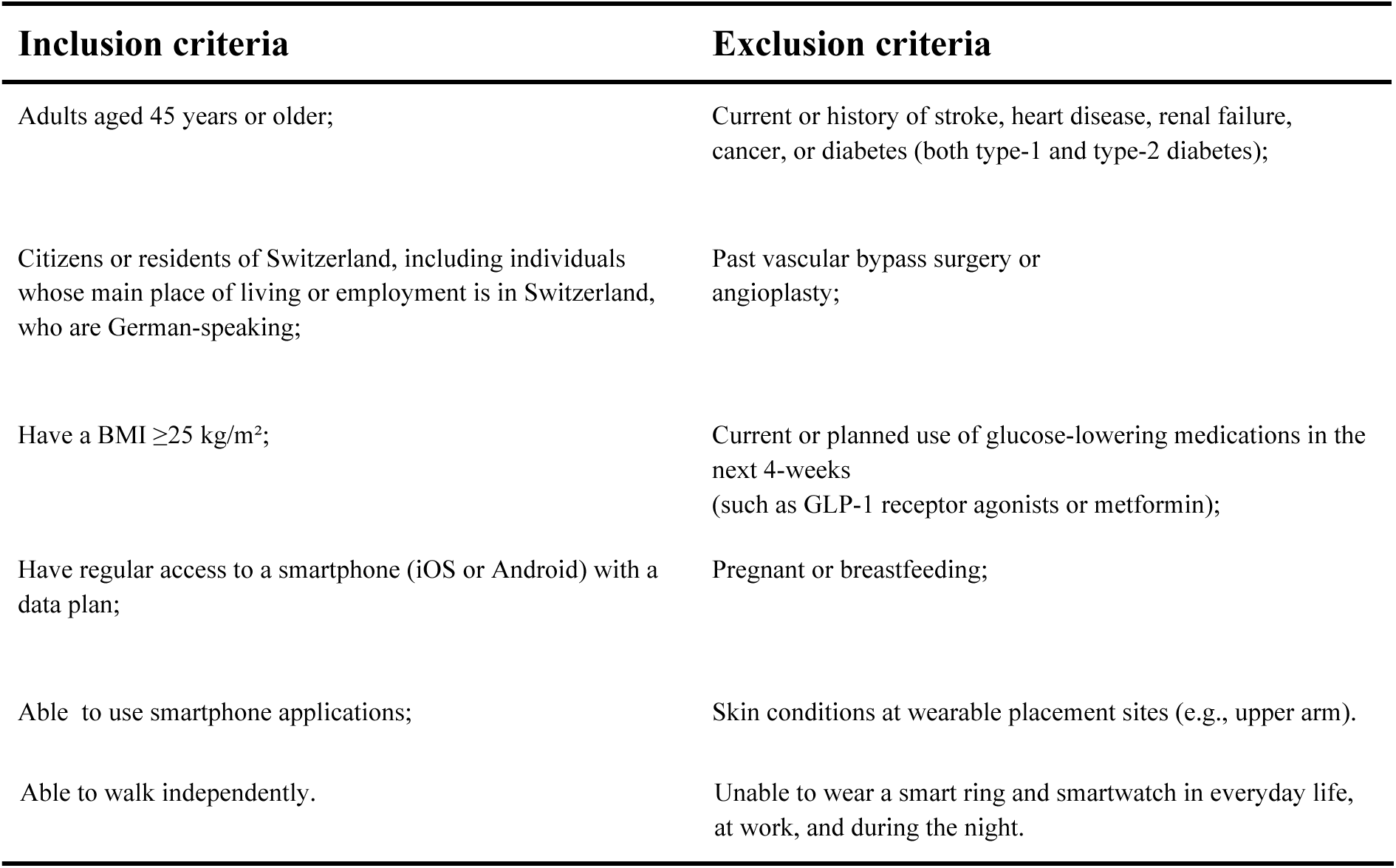
Inclusion and exclusion criteria.

Interested participants are recruited through advertisements via community bulletin boards, post office branches, supermarket blackboards, flyers and in-store commercials, social media (e.g., Facebook ads), as well as through company and institutional networks (e.g., via intranet platforms or newsletters). Participant recruitment is expected to conclude within one year. Individuals interested in participating in the study will be directed to an online survey with the eligibility questionnaire and additional questions from the Swiss T2D risk questionnaire, derived from the Finnish Diabetes Risk Score (see Supplementary Table A3 for questionnaire overview used in this study). (14,42,43). Participants will also be asked to provide personal contact information (phone number and email) to be contacted by the study team for study scheduling and to report their gender to facilitate case-control matching by gender and age. Respondents who are deemed eligible by the study team will be contacted by email to schedule an enrolment visit.

#### Enrolment Visit (T0)

The enrolment visit (T0) is designed to confirm participant eligibility leading to the exclusion of participants with T2D based on a combination of fasting plasma glucose and HbA1c blood measurements. One day prior to the visit, participants will receive an email reminder to fast overnight for at least 8 hours in preparation for the FPG blood test. Once informed consent is obtained, study personnel will review the enrolment questionnaire with the participant by administering each eligibility and T2D risk score question. Participants’ ring sizes and non-dominant arm will be recorded to ensure a proper fit and availability for the finger-worn fitness tracker (Ultrahuman Ring AIR) and the wrist-worn fitness tracker (Ametris Leap), which will be provided at the baseline visit for eligible participants (T1). To check for further eligibility participants will undergo a venous blood draw for HbA1c (ethylenediaminetetraacetic acid tubes) followed by FPG (sodium fluoride tubes) testing by trained medical staff (e.g., a study nurse). Following the two blood tests individuals will be asked to stand on an electronic scale (Withings Body Scan). Waist and hip circumference will also be measured using a measuring tape. Lastly, the following study visits (T1, T2, T3) will be scheduled, depending on their preference, individuals can complete the baseline visit (T1) and intermediate visit (T2) at the study site (HOCH Health in St. Gallen) or at an alternative location, such as their home or workplace. The entire enrolment visit is expected to last approximately 30 minutes.

Based on the blood results (HbA1c and FPG), and self-reported responses to the Swiss T2D risk questionnaire, participants will be allocated to the case or control group using blinded group assignment until follow-up, or excluded as ineligible. Participants with FPG levels between 5.6 and 6.9 mmol/L and/or HbA1c of 5.7-6.4% will be assigned to the case group. Those FPG below 5.6 mmol/L and/or HbA1c of <5.7% will be assigned to the control group. Individuals with valid FPG> 7 mmol/l and HbA1c of >6.4% will be informed of their blood test result and advised to visit a general practitioner for further evaluation.

#### Baseline Visit (T1)

The T1 onboarding visit equips participants with four digital tools: (1) a commercial-grade and a (2) medical-grade fitness tracker, (3) a continuous glucose monitor (CGM), and (4) a study smartphone app. Additionally, self-reported survey data are collected during this visit. See Figure A1 for devices and features collection.

The onboarding process begins with setting up the finger-worn (Ultrahuman Ring Air) and wrist-worn (Ametris formerly Actigraph Leap Watch) (see Figure A1 and Table A2 & for data collected). Each participant will be guided through the setup process, which includes downloading the corresponding app for the consumer-grade device and logging in using their Apple or Google Play account. Following the setup of the fitness trackers, participants will download the study app (MyDataHelps (44)) onto their personal smartphones and the instructions on how to complete survey questions within the app and upload photos of their meals as part of the dietary data collection process will be given after the CGM set-up.

Next, participants will be provided with a CGM device (Abbott FreeStyle Libre Pro iQ), which has been widely used in previous studies(45,46). The CGM will be applied to the non-dominant upper arm and worn continuously for twice 14 days. The blinded CGM needs to be initialized with the CGM Reader (Abbott FreeStyle Libre Pro iQ) to start collecting interstitial blood data every 15 minutes.

After all devices are installed, a full-body photo to assess measurements such as BMI, body fat percentage, and visceral fat will be taken (47). The full-body photo will be taken through the study app on their smartphones. Further, participants will complete a 20-minute online questionnaire via the study app. This questionnaire assesses dietary preferences, typical meal times, and other diet– and well-being-related factors. Responses will be used to customize the timing of meal-tracking notifications and survey prompts to align with each participant’s usual eating patterns. The entire T1 visit, including device setup and questionnaire completion, will take approximately 45 minutes.

#### Remote monitoring period and intermediate visit (T2)

Following the baseline visit, participants will begin a four-week continuous monitoring phase. During this observational period, data on daily diet, physical activity, sleep, body temperature, and stress levels will be continuously collected using the fitness tracker and the study app (see Table A2 for data collection variables). Automated reminders and guidance on device usage, including charging instructions, will be provided to support adherence to the data collection protocol.

Participants will be asked to upload images of all food and beverage consumption three times daily, aligned with their typical meal-time windows (defined as one-hour intervals for breakfast, lunch, and dinner, based on their baseline questionnaire responses). They will also have the option to include annotated manual entries or adjust their food logs during each upload session (see Appendix Figure A1 food tracking upload interface). If a meal has not yet been logged, a reminder notification will be sent at a random time within the relevant one-hour window. If a participant skips a meal, they may select “Skipped Meal” in the app and will be prompted to answer brief follow-up questions about the reason. Snacks consumed between meals can also be optionally recorded using the designated feature in the app, either via image uploads or manual entries. Additionally, participants will be prompted to complete a brief nightly survey in the app to assess mood and stress levels after logging their dinner. To maintain data quality, participants will receive regular reminders to charge their fitness trackers (every other day) and to synchronise the wearable device with its corresponding app if no data has been received in the past 48 hours.

#### Intermediate Visit (T2)

After 14 days of remote monitoring, participants will be invited to attend an intermediate visit within a 7-day window (Day 14-21) to upload and replace the CGM device and upload the wrist-worn fitness tracker data, as the data is stored locally on both devices.

#### Follow-Up On-Site Visit (T3)

The follow-up visit will take place at HOCH Health, where trained medical staff (e.g., study nurses) will collect HbA1c and FPG samples, following the same procedures used during the enrolment (T0) visit. Following blood collection, participants will undergo body composition assessments. They will be asked to stand on the smart scale for anthropometric measurements. Waist and hip circumference will be measured using a standard measuring tape.

Participants will then complete a follow-up questionnaire in the study app that will assess self-reported changes in lifestyle behaviors, adherence to study procedures, perceived usability of the finger– and wrist-worn fitness trackers and attitudes toward digital biomarkers (see Table A3 for questionnaire overview).

At the conclusion of the visit, participants will be debriefed on the study’s progress and compensated based on their adherence to the study protocols. Participants will also be asked to return their CGM device and fitness trackers. Finally, they will be given the option to either retain or delete the study data stored on their personal smartphones. Moreover, they can opt in or out of being contacted during the data retention period for follow-up studies. Finally, participants will be reimbursed with up to 170 CHF according to their completion of visits and adherence to meal image uploads, and a report of their data collected will be provided a couple of weeks after their follow-up visit. During data collection, data confidentiality will be ensured and all data will be retained for 10 years in accordance with institutional and ethical guidelines.

### Study assessments

The multimodal data collection variables during the study are described in Appendix, Figure A2; please refer to Appendix 1, Table A1, for the frequency of feature measurements. This study poses minimal physical risks to participants, primarily related to blood sampling and the use of CGM. Participants will be informed in the event of adverse events and advised to discontinue participation or, if clinical diagnostics reach the T2D threshold, to reach out to their GP. To address any potential risks of data privacy, all participant data will be handled in strict compliance with data protection regulations, including the Swiss Federal Act on Data Protection and the EU General Data Protection Regulation. Data will be pseudonymized before analysis and stored on encrypted servers with access restricted to authorized study personnel only. Participants will be promptly notified in the event of any data leakage or breach involving their personal information. Issues that arise will be collected and reported according to best practices.

## ANALYSIS

### Statistical Analysis

#### Sample size

As this is a study for testing the feasibility of a prediction model, prediction models are often developed with no sample size calculator, yet recommendations entail to ensure at least 10 events for each predictor parameter and Riley et al.’s sample size using the *pmsamplesize* R package(48), following Riley et al.’s framework for developing clinical prediction models with binary outcomes. The calculation was based on five predictor variables (including lifestyle and demographic factors), a shrinkage factor of 0.9, and an expected Nagelkerke’s R² of 0.3(49), which is consistent with similar studies using physiological and demographic predictors. This approach yielded three sample size estimates corresponding to different criteria: (1) ensuring adequate shrinkage to limit overfitting and improve model generalisability (n = 174); (2) ensuring small absolute differences between the apparent and adjusted Nagelkerke’s R² (n = 115); and (3) ensuring a precise estimation of the overall outcome risk (n = 385). Considering the primary feasibility goal of the current project and budgetary constraints, we selected a target sample size of 200 participants (100 controls and 100 individuals with prediabetes), accounting for a dropout rate of 15%. The reason for withdrawal from the study will be documented and if needed, additional participants will be recruited.

#### Data Analysis

Data quality and data management checks will be conducted throughout the study in accordance with established standards, and any inconsistencies will be documented to support subsequent data cleaning. Following data cleaning and completeness checks, statistical analyses will be conducted using R (version 4.3.1 or newer) and Python (version 3.11 or newer). Non-responders and missing data will be analysed and reported according to best practices, with appropriate imputation methods applied where needed. Descriptive statistics (means, SDs, frequencies) will be used to summarise baseline demographics, anthropometric data, and wearable-derived lifestyle metrics.

The primary outcome, classification of prediabetes based on wearable-derived lifestyle metrics and baseline demographics, will be analysed using machine learning classification models, including logistic regression, random forests, and gradient boosting. Predictive performance will be evaluated using area under the curve (AUC), accuracy, sensitivity, and specificity. In parallel, descriptive analysis will be performed.

Secondary analyses will include multivariate regression, machine learning and network analysis models to explore associations between lifestyle variables (e.g., HRV, stress, nutrition), continuous glucose outcomes (e.g., postprandial spikes, time-in-range) and metabolic subgroups. Moreover, metabolic subgroup comparison will be assessed using χ² tests for categorical variables and t-tests or ANOVA for continuous variables. Adherence to image-based meal tracking and effects of dynamic notifications will be analysed using linear mixed models (39). A *p*-value of <0.05 will be considered statistically significant.

## DISCUSSION

The Glow Up study is among the first observational digital biomarker studies to investigate lifestyle patterns among normoglycemic individuals and individuals with prediabetes by creating a richly digitally phenotypes, multimodal dataset in an at-risk population under real-world conditions over 4 weeks in Switzerland. The study integrates four complementary data modalities: (1) continuous lifestyle data derived from commercial and medical-grade fitness trackers, including HR, HRV, sleep, physical activity, and body temperature, as well as smartphone app data with stress indicators and image-based meal tracking; (2) CGM data capturing high-resolution interstitial glucose dynamics; (3) clinical blood-based biomarkers, including HbA1c and FPG; and (4) anthropometric and body composition measures obtained via a smart scale. The overarching aim of the study is to test the feasibility of a digital biomarker for prediabetes screening and lifestyle phenotyping in individuals at risk for T2D.

A key strength of the Glow Up study is the combination of multiple, complementary data streams collected over 4 weeks. The study uniquely integrates consumer-grade devices (smart ring, smartphone-based meal tracking, smart scale) with medical-grade tools and measurements (CGM, smartwatch, and clinical blood biomarkers), allowing reliability comparison with established clinical standards. This multimodal design enables assessment of unique combinations of burden and scalability of lifestyle-derived metrics for prediabetes prediction. The longitudinal and multimodal structure of the study also creates potential for the application of time-series foundation models (50–53). To our knowledge, none of the previously published studies (54–65) in digital phenotyping use the same combination of granularity and data modalities as Glow Up. Previous studies combine clinical tests and lifestyle monitoring, such as the AI-READI dataset (66), Human Phenotype Project (66,67) or COBRA Study (64), are large-scale cohorts with an abundance of biobank, molecular, and clinical tests, or smaller datasets, such as the BIG IDEAs Lab Glycaemic Variability and Wearable Device Data (55,68) with 16 study participants or the Stanford CGMDB (63,69) with 55 study participants. Those studies include less granular and shorter-duration lifestyle and behavioral data than Glow UP, which mainly focuses on digital tool data collection. Further studies also focus on digital tool data collection (57,61); however, they include no clinical tests. Glow Up extends prior research in Switzerland from the Food & You study by Héritier et al. (2023) (57) by including individuals with prediabetes in addition to normoglycemic individuals, and by adding clinical laboratory measures for gold-standard prediabetes outcome assessment and anthropometrics.

Glow Up’s glucose blood biomarker measurements extend another important strength, namely the inclusion of both baseline and follow-up blood measurements of FPG and HbA1c. Although the study is observational, the repeated blood measurement assessments enhance internal validity and allow observation of within-person trends in the impact of the digital tools used during the study, including the Hawthorne effect (70), although not eliminating it. This is particularly relevant as changes in lifestyle could already lead to changes in FPG within 4 weeks (71). The case-control design, with age– and sex-matched normoglycemic and prediabetic participants, further supports meaningful group comparisons while minimizing confounding.

Besides the lack of a control group, the observational period of approximately four weeks, although longer than in other studies in a similar context (59,61,72), is short relative to the biological time frame of two to three months for HbA1c (12). Nevertheless, a four-week monitoring period is sufficient to capture short-term blood glucose dynamics, including fasting plasma glucose and CGM-derived measurements of glycaemic variability.

Recruitment was structured in a diverse way (e.g., outreach through different institutions, in-person recruitment at post offices, and social media) to reach a broad population. However, due to the involvement of university and medical networks, the study may be prone to selection bias toward individuals who are more interested in digital tools and health.

Although multimodal data were collected, measures of gut health (57) and metabolic sub phenotypes (63) (i.e. insulin suppression test, lipid panel) were not recorded due to infrastructural and cost constraints. This limits the ability to capture additional metabolic mechanisms underlying glucose regulation.

Future research should build on the Glow Up study by extending the observational period, incorporating long-term longitudinal follow-up, adding further measurements such as metabolic sub phenotype, and potentially evolving into a prospective cohort study. While the primary objective of Glow Up is the development of a digital biomarker for prediabetes screening and the investigation of digital phenotypes, subsequent research phases should focus on translating these biomarkers into actionable interventions, such as just-in-time adaptive interventions. These may include personalized, data-driven lifestyle interventions delivered through artificial intelligence–based health coaching by implementing for example time series foundation models.

## Data Availability

All data produced in the present study are available upon reasonable request to the authors

## Acknowledgements

The authors would like to thank all study investigators, study personnel, affiliated staff, and partner networks for their valuable contributions to the conduct of the study and their support with participant recruitment. We are particularly grateful to the study nurse Victoria Schlenker and the master’s thesis student Zoe Przygienda for their dedicated assistance with data collection.

The Ultrahuman Ring AIR devices were provided by Ultrahuman Healthcare Pvt Ltd, Ametris LLC, and Withings France SA.

## Author contributions

The study concept and design was conceived by VB, MF, TK, and MJ. BW, MB, QJ and SB assisted in refining study design. VB, MF and QJ are responsible for data collection. Analyses will be conducted by VB, MF, QJ and MJ. VB prepared the first draft of the manuscript. All authors critically revised the manuscript and approved the submitted version. AI tools were used to generate the images in Figure A2 and to proofread the text of this article.

## Competing interest declaration

**Ethical approval:** The study has received ethics approval from the Ethics Committee of Eastern Switzerland (BASEC ID.: 2025-00972).

[**Trial registration:** NCT07373418]

**Role of study sponsors:** This study is a Sponsor-Investigator study. PIs include Prof. Dr. Tobias Kowatsch and Dr. Mia Jovanova.

## Funding

VB, MF, QJ, TK and MJ are affiliated with the Centre for Digital Health Interventions, a joint initiative of the Institute for Implementation Science in Health Care, University of Zurich, the Department of Management, Technology, and Economics at ETH Zurich, and the Institute of Technology Management and School of Medicine at the University of St.Gallen, Centre for Digital Health Interventions is funded in part by Mavie Next, an Austrian health care provider, CSS, a Swiss health insurer, and MTIP, a growth equity firm. TK was also a co-founder of Pathmate Technologies, a university spin-off company that creates and delivers digital clinical pathways. However, Mavie Next, CSS, MTIP or Pathmate Technologies were not involved in this protocol. The other authors declare no further competing interests.

Devices were supplied in kind by Ultrahuman Healthcare Pvt Ltd, Withings France SA and Ametris LLC, who had no role in study design, data collection, analysis, or interpretation.

## Patient and public involvement

During pilot testing, we developed and fine-tuned study procedures in the target study population and a broader population to ensure appropriate procedures, streamlined processes and respectful handling. The results of each participant’s own assessments will be disseminated to them, and general Glow Up results will be disseminated to the general public as appropriate.

### Other declarations: Data sharing

No additional data are available for this study protocol.

### Provenance and peer review

Not commissioned, externally peer reviewed.

